# Exploring Local Health Knowledge and Access: Focus Group Findings from Community Health Workers in Pader District, Uganda

**DOI:** 10.1101/2025.09.11.25335608

**Authors:** Brett R. Albee, Atiya Patrick Kasagara, Denis Otema, Olanya Denish, Isaac V. Faustino, Dhatri Abeyaratne, Shayna D. Cunningham, Rogie Royce Carandang, Felix Bongomin, Daniel S. Ebbs

## Abstract

In Northern Uganda, communities continue to face significant health challenges driven by a legacy of conflict, poverty, and structural health system challenges. Community health workers (CHW) play a critical role in rural districts like Pader, where access to formal care remains limited. This study explores CHWs’ perspectives on community health needs, barriers to care, and priorities for strengthening CHW-led service delivery. Six focus group discussions (7-9 participants each) were conducted with 46 CHWs from across Pader District using a semi-structured topic guide. The guide explored eight key domains: general health, health maintenance, modern versus traditional medicine, nutrition, maternal and infant health, environmental factors, malaria practices, and feedback for the CHW program. CHWs identified malaria, maternal and child health complications, and a rising burden of non-communicable diseases as key community concerns. Major barriers to care included long distances to health facilities, drug stockouts, and inadequate transportation. Although communities had strong awareness of disease symptoms and prevention strategies, financial hardship and service limitations undermined preventive practices and timely care-seeking behavior. CHWs emphasized their growing role as trusted frontline providers but highlighted the need for tools, transportation, ongoing training, supportive supervision, and formal recognition to fulfill their responsibilities effectively. CHWs in Pader District navigate complex structural and resource constraints while serving as trusted liaisons between communities and the formal health system. Their insights point to actionable strategies, such as improved supply chains, transportation, training, and inclusion in program planning, that are vital for building equitable and effective community health systems in post-conflict settings.

## Introduction

Rural and post-conflict regions across sub-Saharan Africa continue to face entrenched health disparities driven by poverty, geographic isolation, and the long-term impacts of political instability. Northern Uganda exemplifies these challenges, particularly in districts like Pader, which report some of the country’s poorest health outcomes despite national progress [1].

Fragile health systems, limited infrastructure, and under-resourced facilities continue to constrain service delivery and undermine health equity in these regions. Decades of armed conflict, most notably the insurgency led by the Lord’s Resistance Army, devastated Northern Uganda’s health system, displacing over 1.8 million people and disrupting access to basic services [2]. While peace has been restored since the mid-2000s, the region remains burdened by chronic poverty, underdeveloped infrastructure, and institutional mistrust.

Community health workers (CHWs) play an essential role in extending the reach of the health system in rural and post-conflict settings. As trusted members of their communities, they serve as critical liaisons between households and formal health services, helping to bridge gaps in access and rebuild trust in a health system still recovering from the effects of conflict and chronic under-resourcing [3]. However, CHWs often operate in highly constrained environments, with chronic supply shortages, limited training, and a lack of formal recognition [3]. Despite their importance, little is known about how CHWs in post-conflict settings perceive the health concerns of their communities, the systemic constraints they face, or the supports they require to deliver effective care. Existing national health surveys, such as the Uganda Demographic and Health Survey, provide important district-level data, but are limited in their ability to capture the lived realities at the village level where health decisions are made and barriers are most acute [4]. This gap in understanding limits the ability of health programs to adapt to local needs.

This study addresses this gap by exploring the experiences and perspectives of CHWs in Pader District, Northern Uganda to (1) identify local health priorities, (2) examine barriers to care and health-seeking behaviors, and (3) generate CHW-informed recommendations to strengthen CHW-led service delivery. Findings will inform the design of a CHW-led community health assessment tool and offer actionable insights for strengthening community health systems in similarly under-resourced, post-conflict settings.

## Materials and Methods

### Study Design and Setting

This qualitative descriptive study was guided by the interpretive paradigm and conducted in collaboration with the Laro Kwo Project, a community-based health initiative operating in Pader District, Northern Uganda. The Laro Kwo Project was established in 2016 through a partnership between local leaders, Ugandan health professionals, and international collaborators, with the aim of strengthening grassroots healthcare delivery in post-conflict Northern Uganda. The program trains and supports CHWs to deliver basic health education, screening, and referral services at the village level [6].

Focus group discussions were conducted in six sub-counties where the Laro Kwo Project is active: Awere, Pader Town Council, Pukor, Puranga, Kilak, and Pajule. At the time of data collection, the program had trained over 150 CHWs, who were distributed across these six sub-counties. This study forms part of a larger mixed-methods evaluation of the Laro Kwo Project. The interpretive paradigm allowed for an in-depth understanding of CHWs roles and experiences within the broader health system and local community context.

### Study Population

CHWs were selected through purposive sampling to ensure diverse representation across gender, age, program experience, and geographic location. Recruitment was facilitated by the program coordinator and CHW leaders from each sub-county. Each sub-county represents a group of CHWs with differing years of experience, and diversity in geographic area. CHWs were included in the study if they were over 18 years of age and had been active participants in the Laro Kwo Project for at least 6 months.

### Data Collection

Six focus group discussions (FGDs) with a total of 46 participants were conducted between November 20 and 28, 2024. One FGD was held in each of the six sub-counties where the Laro Kwo Project operates. Participants in each FGD were drawn from the same sub-county to ensure localized perspectives and to facilitate open discussion among familiar peers. Each group included 7 to 9 participants and sessions lasted between 90 and 120 minutes.

The FGD guide was developed collaboratively by the research team in partnership with Ugandan collaborators, including CHWs and local leaders, to ensure cultural relevance and alignment with community priorities. The guide included open-ended questions across eight domains: general health, health maintenance, modern versus traditional medicine, nutrition, maternal and infant health, environmental factors, malaria knowledge and practices, and program recommendations (Appendix 1).

The FGDs were co-facilitated by the co-principal investigators—one from the United States and one from Northern Uganda—alongside the Laro Kwo Project’s local research and program coordinator, who played a key role in planning, logistics, and translation. Discussions were moderated in English, with real-time translation into Acholi when needed, to ensure clarity and comfort for all participants. Facilitators took detailed field notes, capturing both direct quotations and contextual observations.

### Data Analysis

Data were analyzed using a interpretative phenomenological analysis (IPA) [7]. Two members of the research team, the co-principal investigators (BA and APK), independently reviewed and open-coded notes from the FGDs to create the initial codebook. We first assigned codes to meaningful phrases and then assigned text with similar meaning to the same codes. Next, we collected similar codes into different categories where we assigned themes and sub-themes. The codebook was refined through team discussions to resolve any discrepancies. Themes were developed inductively and organized around both the domains from the guide and themes that arose from the discussion participants. Illustrative quotes were selected to represent key findings. The study methods adhered to the Consolidated Criteria for Reporting Qualitative Studies (COREQ) [8]. A complete checklist is included (Appendix 2).

### Ethical Considerations

This study was approved by the Yale University Institutional Review Board (IRB#2000038006) and the Gulu University Research Ethics Committee (GUREC-2024-854), and also received formal authorization from the Pader District Government Health Department. CHWs were enrolled in focus groups from 11/20/2024 to 11/28/2024. Verbal informed consent was obtained from all participants after receiving consent forms and expressing comprehension of study. A program coordinator and co-PI documented consent after acknowledging comprehension of study in either Acholi or English.

## Results

Table 1 summarizes participant characteristics. The participants ranged in age from 29 to 58 years old, and spanned the entire lifetime of the Laro Kwo Project; one group included the first CHWs to join the program in 2016, and another group included the most recent CHWs to join just six months prior. Almost one third of all CHWs within the Laro Kwo Project were represented in the focus groups, with 46 participating out of 150 total CHWs.

**Table 1.** Focus Group Participant Characteristics.

| Focus group setting | Number of Participants | Number of Males (%) | Average Age (years) | Years of experience in Laro Kwo Project |
| --- | --- | --- | --- | --- |
| Puranga | 7 | 6 (86) | 45.6 | 5 |
| Awere | 7 | 7 (100) | 41.7 | 6 |
| Kilak | 8 | 6 (75) | 51.4 | 8 |
| Pajule | 9 | 7 (78) | 37.8 | 0.5 |
| Pader Town Council | 7 | 3 (43) | 43.4 | 3 |
| Pukor | 8 | 6 (75) | 38.3 | 3 |

Participants provided rich and multidimensional accounts of health dynamics within their communities, encompassing both clinical priorities and the sociocultural contexts that shape care-seeking practices. IPA analysis yielded key insights into the social and structural determinants of health in the region, as well as the evolving roles, experiences, and challenges faced by CHWs in bridging community needs with formal health services (Table 2). The following themes emerged: community health concerns, healthcare quality and access, and knowledge versus practice.

**Table 2.**
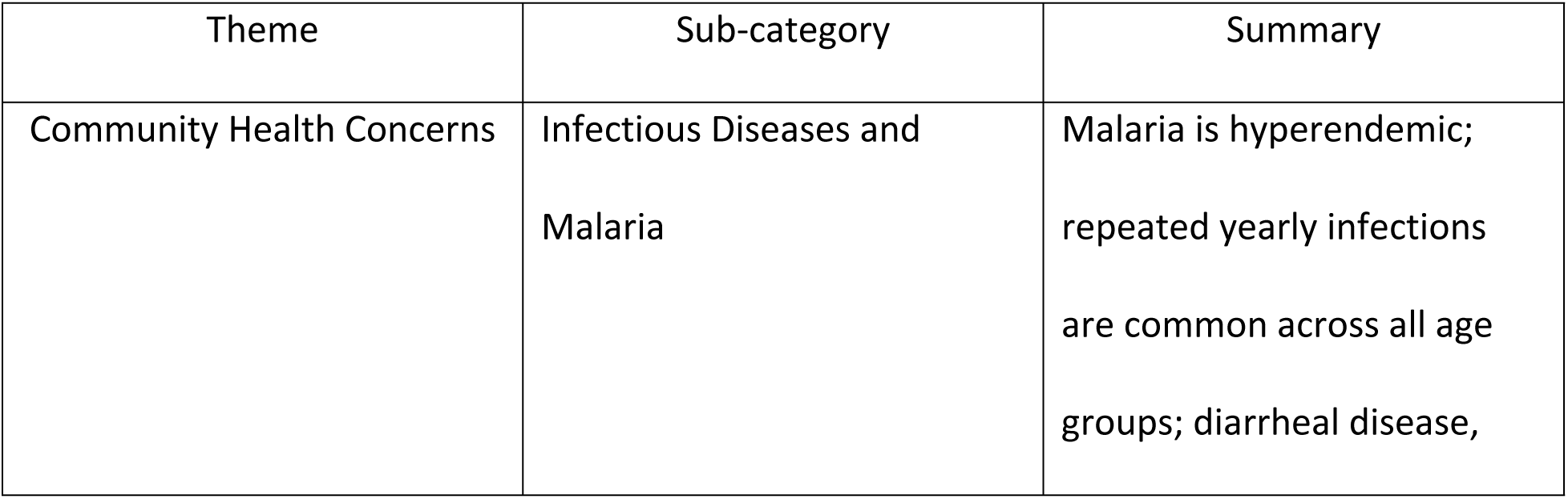
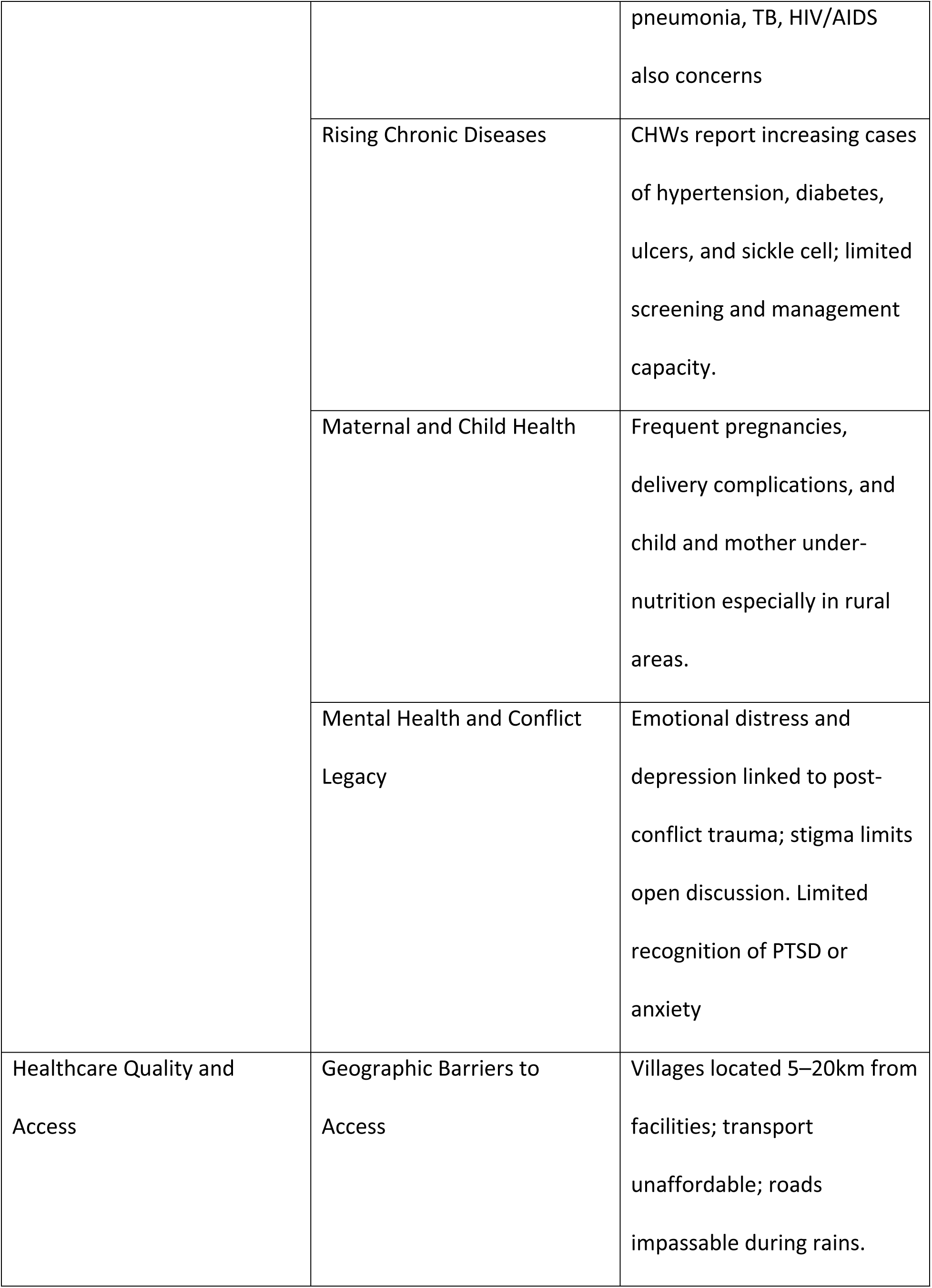
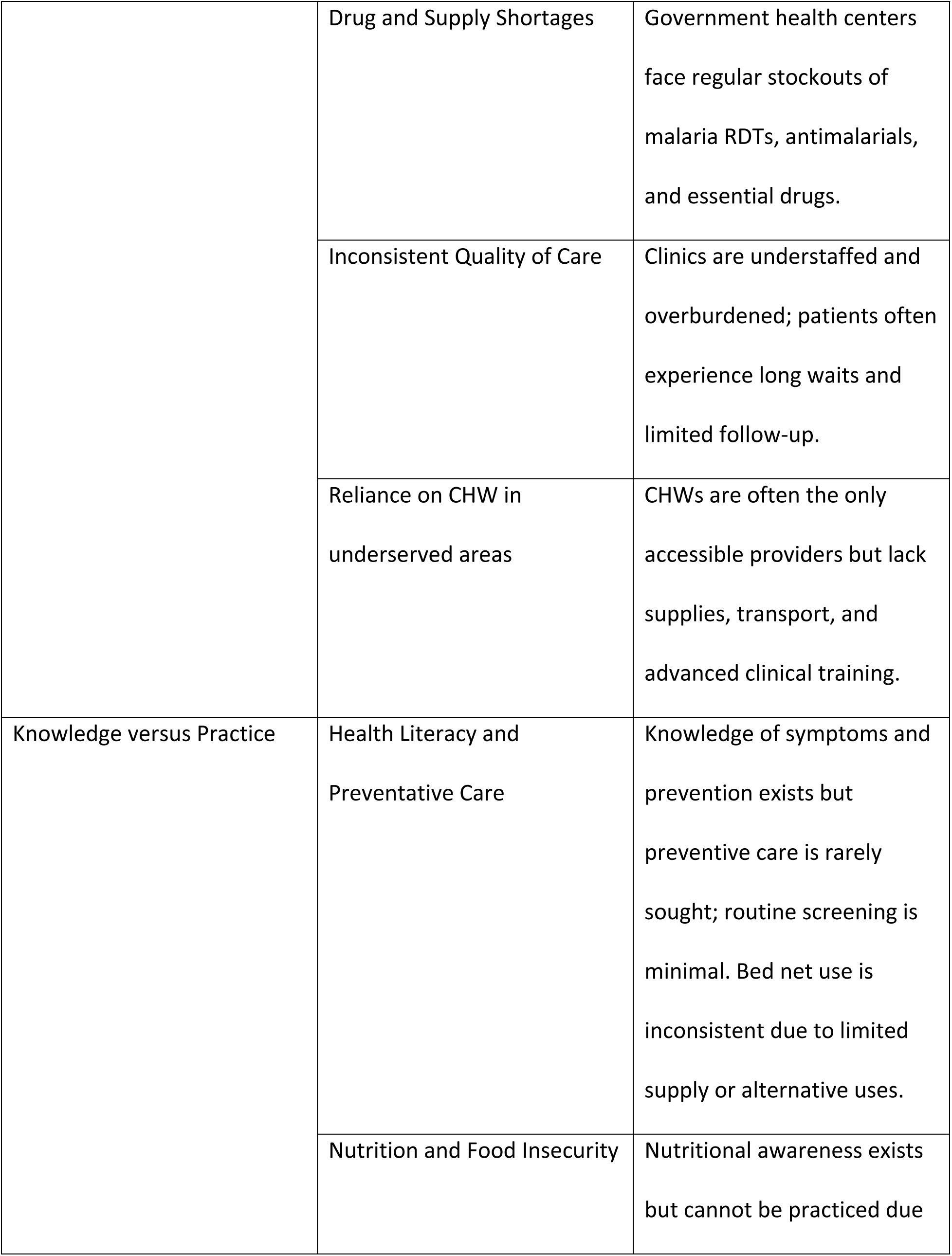
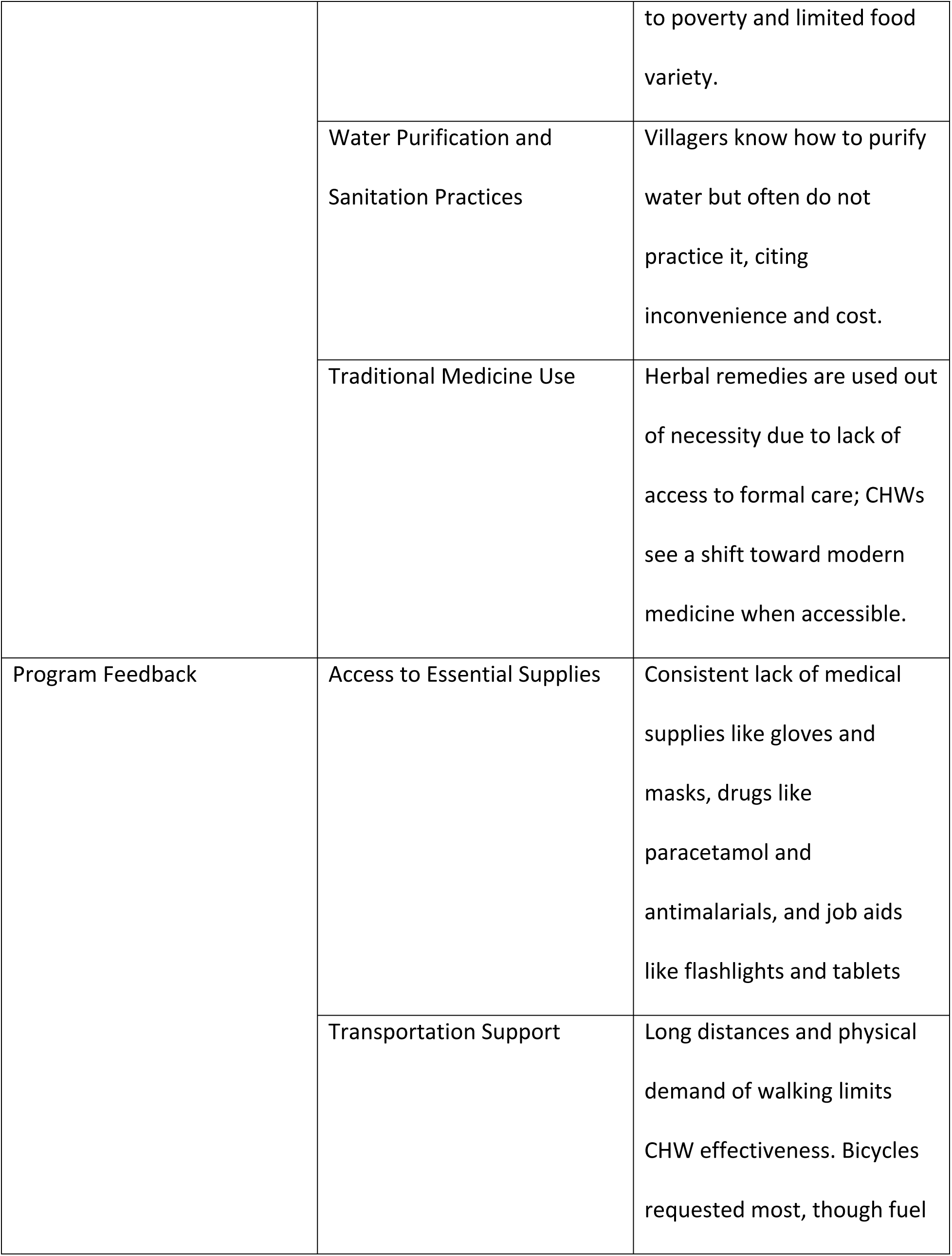
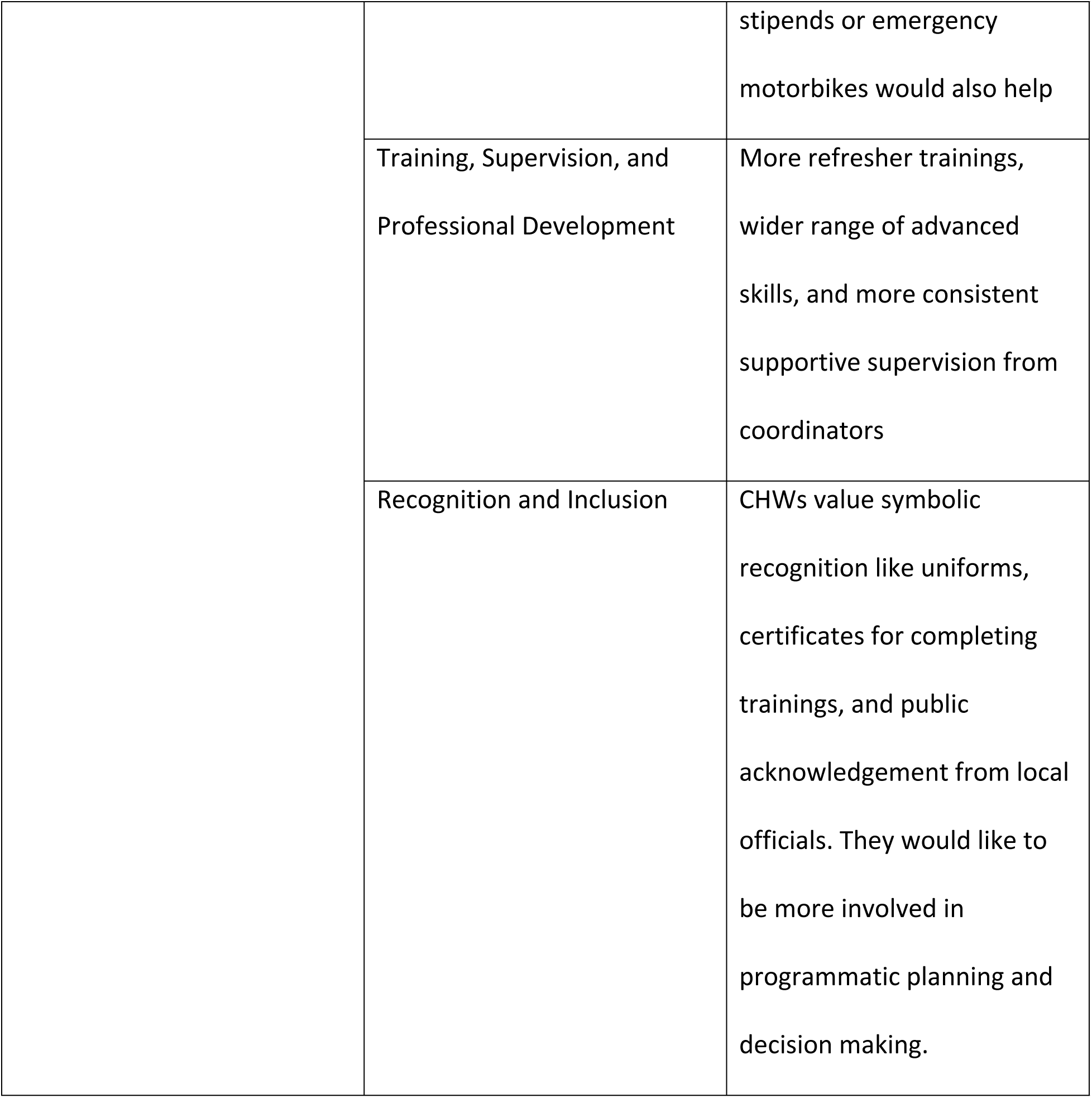
Main themes from FGD analysis.

| Theme | Sub-category | Summary |
| --- | --- | --- |
| Community Health Concerns | Infectious Diseases and<br>Malaria | Malaria is hyperendemic;<br>repeated yearly infections<br>are common across all age<br>groups; diarrheal disease, |
|  |  | pneumonia, TB, HIV/AIDS<br>also concerns |
|  | Rising Chronic Diseases | CHWs report increasing cases of hypertension, diabetes, ulcers, and sickle cell; limited screening and management capacity. |
|  | Maternal and Child Health | Frequent pregnancies, delivery complications, and child and mother under-nutrition especially in rural areas. |
|  | Mental Health and Conflict Legacy | Emotional distress and depression linked to post-conflict trauma; stigma limits open discussion. Limited recognition of PTSD or anxiety |
| Healthcare Quality and Access | Geographic Barriers to Access | Villages located 5–20km from facilities; transport unaffordable; roads impassable during rains. |
|  | Drug and Supply Shortages | Government health centers face regular stockouts of malaria RDTs, antimalarials, and essential drugs. |
|  | Inconsistent Quality of Care | Clinics are understaffed and overburdened; patients often experience long waits and limited follow-up. |
|  | Reliance on CHW in underserved areas | CHWs are often the only accessible providers but lack supplies, transport, and advanced clinical training. |
| Knowledge versus Practice | Health Literacy and Preventative Care | Knowledge of symptoms and prevention exists but preventive care is rarely sought; routine screening is minimal. Bed net use is inconsistent due to limited supply or alternative uses. |
|  | Nutrition and Food Insecurity | Nutritional awareness exists but cannot be practiced due |
|  |  | to poverty and limited food variety. |
|  | Water Purification and Sanitation Practices | Villagers know how to purify water but often do not practice it, citing inconvenience and cost. |
|  | Traditional Medicine Use | Herbal remedies are used out of necessity due to lack of access to formal care; CHWs see a shift toward modern medicine when accessible. |
| Program Feedback | Access to Essential Supplies | Consistent lack of medical supplies like gloves and masks, drugs like paracetamol and antimalarials, and job aids like flashlights and tablets |
|  | Transportation Support | Long distances and physical demand of walking limits CHW effectiveness. Bicycles requested most, though fuel |
|  |  | stipends or emergency motorbikes would also help |
|  | Training, Supervision, and Professional Development | More refresher trainings, wider range of advanced skills, and more consistent supportive supervision from coordinators |
|  | Recognition and Inclusion | CHWs value symbolic recognition like uniforms, certificates for completing trainings, and public acknowledgement from local officials. They would like to be more involved in programmatic planning and decision making. |

### Community Health Concerns

CHWs across all focus groups identified a wide range of health conditions affecting their communities, with malaria consistently described as the most pervasive and urgent concern. Participants reported that nearly every household experiences multiple malaria episodes each year, with children under five and pregnant women particularly vulnerable to severe illness. One male CHW with 8 years of experience noted, “*Everyone here has had malaria many times, even in the same year. It is just normal now*” (Kilak). Seasonal variation was noted, with increased cases during the rainy season.

Beyond malaria, CHWs identified diarrheal diseases, pneumonia, tuberculosis, HIV, and intestinal worms as common challenges. Maternal and child health concerns, including frequent pregnancies, malnutrition, and complications during childbirth, were particularly acute in remote areas. Noncommunicable diseases such as hypertension, diabetes, epilepsy, and cervical cancer were also mentioned with increasing frequency, though CHWs emphasized the lack of screening and awareness. Mental health issues, especially depression and emotional distress linked to the long-term effects of conflict, were noted as widespread but rarely addressed due to stigma and limited access to specialized care. Across all conditions, CHWs pointed to delayed care-seeking and inadequate access to health services as key contributors to poor outcomes. The burden of disease, they stressed, was compounded by systemic and structural barriers that are addressed in subsequent themes.

### Healthcare quality and access

Across all focus groups, CHWs described significant and persistent barriers to accessing timely and appropriate healthcare. These barriers were largely structural, stemming from geographic isolation, inadequate transportation, and chronically under-resourced health facilities. CHWs described a context in which health facilities are typically located 5-7 kilometers from most villages, with some communities situated up to 20 kilometers away. Hospitals, which are essential for emergency and advanced care, are often located even farther, making access nearly impossible for many, particularly during urgent situations. Participants emphasized that many individuals are simply too ill, too impoverished, or both, to make such journeys.

Transportation options are limited, motorbikes and bicycles are scarce, and most families cannot afford the cost of fuel, transport fares, or even mobile phone airtime to request assistance. While an ambulance system was recently introduced and viewed positively by CHWs, it still requires patients to cover fuel costs, which presents a barrier for many. Poor road infrastructure further compounds the problem, with unpaved, unlit roads that become impassable during the rainy season.

Access to treatment is similarly constrained. Government health centers frequently run out of artemether-lumefantrine, the recommended first-line therapy, and private pharmacies, though better stocked, are unaffordable for many. CHWs expressed concern about declining drug efficacy and poor adherence, with some patients taking incomplete courses or saving doses for future illness. One CHW, who joined the LKP 6 years prior, noted, “People are taking the drugs, but they don’t always get better. We think the medicine is not working like it used to” (Awere).

Reaching a facility, however, does not guarantee quality care. Government-run health centers offer services at no cost, but they are often under-resourced and understaffed. Essential medications are frequently out of stock, and staff shortages can lead to long wait times and overburdened healthcare workers, potentially compromising the quality of patient interactions. CHWs noted that drug shipments typically arrive only once per quarter and are rapidly depleted. In contrast, private clinics and pharmacies tend to have more reliable supplies, but their services are financially out of reach for most community members. As a result, patients may visit multiple facilities in search of treatment or ration medications to make them last, a strategy that, while understandable, contributes to poor health outcomes and may exacerbate problems such as antimicrobial resistance.

### Knowledge versus practice

CHWs reported that health knowledge within communities varied widely, with limited awareness of preventive care and widespread delays in seeking formal healthcare services. Many community members reportedly wait until illness becomes severe before visiting a clinic, often bypassing early intervention opportunities. One female CHW explained, “*People only go [to the clinic] when it is already bad. They don’t check their health unless something is wrong*”(Pader Town Council). Preventive services, such as blood pressure monitoring or cancer screening, are rarely sought or available outside district hospitals.

Even when individuals possess some knowledge about disease prevention, economic hardship and systemic limitations often prevent them from acting on this knowledge. CHWs described a consistent disconnect between knowledge and practice. For example, while many families are aware of the importance of nutrition, balanced diets are often unattainable due to food insecurity and poverty. “*They know they should eat a balanced diet,”* one CHW noted*, “but they just eat what is available*”(Pajule). Diets are dominated by inexpensive staples such as maize and beans, with limited access to animal protein or fresh produce. Seasonal shortages further restrict food availability, and some households intentionally reduce their food intake in order to sell more crops to cover school fees or healthcare costs.

Despite high community awareness of malaria symptoms and the importance of early treatment, CHWs described frequent barriers to effective prevention and care. Insecticide-treated bed nets are often insufficient in quantity, distributed infrequently, or repurposed for other uses such as fishing. Proper use and installation are inconsistent, with minimal guidance provided. Some CHWs also reported that discomfort caused by treated nets, such as headaches, discourages regular use.

Similar gaps were observed in water purification and hygiene practices. While many villagers understand the importance of purifying drinking water, they often view the process as unnecessary or burdensome. CHWs explained that boiled or treated water is seen as an extra step rather than a necessity. As one participant put it, “*People know how to purify water, but they say it’s a waste of time*”(Puranga). Most households rely on wells, boreholes, or untreated surface water, with piped water available only in towns and bottled water largely unaffordable.

Traditional medicine use was widespread, often driven by necessity rather than cultural preference. CHWs described the use of herbal remedies such as blackjack flower, neem leaves, and mango bark to treat common ailments like malaria, gastrointestinal issues, and wounds. While some CHWs acknowledged their potential therapeutic value, they also expressed concern that these practices can delay access to formal care or result in adverse effects when used improperly. Nonetheless, participants noted a gradual shift toward biomedical care, especially when CHWs are able to offer guidance and referral. As trusted figures within their communities, CHWs increasingly serve as informal first points of contact for basic care and health education. Many reported being approached for advice, blood pressure checks, and symptom interpretation, even though they often lack the training or resources to provide direct treatment. Another CHW explained how some people treat them as doctors, even though their main responsibility is referring them to clinics.

### Feedback for Laro Kwo Project

CHWs across all focus groups shared detailed, experience-based recommendations for strengthening the Laro Kwo Project and enhancing their ability to serve their communities effectively. Their feedback emphasized the need for both material support and systems-level improvements that reflect their essential role within the health system. These recommendations were grounded in the daily challenges CHWs face and reflect both material needs and systemic gaps that limit their effectiveness. A common and urgent concern was the lack of access to essential medical supplies. CHWs described being expected to deliver care and assess patients without having basic resources such as malaria rapid diagnostic tests, antimalarials, paracetamol, amoxicillin, and zinc. These tools were consistently framed not as supplementary aids, but as core necessities for fulfilling their roles. Several CHWs also emphasized the need for basic equipment such as first aid kits, disposable gloves, and headlamps, particularly for nighttime visits or emergencies. To support documentation and data management, many CHWs requested the provision of waterproof storage bags for paper records, which are frequently damaged during the rainy season. Some also proposed more innovative solutions, including equipping CHWs with tablets or mobile phones preloaded with reference materials, reporting templates, and referral tracking tools. Given the increasing access to smartphones and mobile data in the region, CHWs viewed digital solutions as both feasible and transformative.

Transportation was another major theme. CHWs working in remote areas described walking for hours each day to visit households, accompany patients to health centers, or retrieve supplies. The absence of reliable transport severely limited their reach and contributed to physical exhaustion and burnout. Bicycles were the most frequently requested form of support, although some also proposed fuel stipends or emergency motorbike transport options for urgent referrals.

Training and professional development were cited as critical to both the quality of care and CHW motivation. Participants called for more regular refresher trainings on foundational skills, such as identifying malaria danger signs and measuring vital signs, as well as expanded instruction in areas like maternal and child health, nutrition, mental health, and emergency care. Several CHWs expressed a desire to participate more actively in outreach and education initiatives and requested training in public speaking, recordkeeping, and community engagement strategies. This desire for growth was coupled with calls for stronger, more consistent supervision. Many CHWs reported infrequent contact with program staff and described a sense of isolation from program leadership. They advocated for monthly supervisory visits, quarterly review meetings, and more open channels of communication. A female CHW from Pukor shared, “*Sometimes it feels like we are forgotten. We need someone to come and ask how we are doing*.” Participants emphasized that supervision should be collaborative and supportive, offering guidance, feedback, and encouragement rather than strict oversight. Recognition, both financial and symbolic, emerged as a powerful theme. While CHWs acknowledged the limitations of program budgets, many advocated for more predictable stipend disbursements, subsidized healthcare for themselves and their families, or performance-based incentives. Others stressed the value of symbolic recognition in building legitimacy and community respect. Uniforms, certificates, or public acknowledgment at community gatherings were cited as low-cost but highly meaningful gestures. As another participant explained, “*Even just a uniform shows people that we are real health workers. It builds respect*”(Awere).

Finally, CHWs expressed a strong desire for further involvement in program design and implementation. They emphasized that their intimate knowledge of community dynamics, barriers, and behaviors positions them as vital partners in shaping interventions that are both feasible and culturally appropriate. Participants called for consultative meetings, feedback loops, and opportunities to contribute to decision-making. This inclusion, they argued, would foster greater accountability, strengthen motivation, and ultimately improve the impact of the program.

## Discussion

This study explored the experiences and perspectives of CHWs across Pader District, Northern Uganda, with the aim of understanding community health priorities, barriers to care, and opportunities to strengthen CHW-led service delivery. The findings reveal a landscape marked by high disease burden, entrenched structural barriers, and persistent resource limitations, but also one in which CHWs are trusted, committed, and increasingly positioned as central actors in advancing community health [9].

Malaria is hyperendemic in the region and remains the leading cause of morbidity and mortality, especially among children under five and pregnant women, with CHWs describing near-universal exposure across all age groups and repeated infections throughout the year [4]. While community awareness of malaria symptoms and the need for early treatment was high, prevention and treatment remain compromised by inconsistent access to bed nets, stockouts of antimalarials, and emerging concerns about treatment efficacy. These findings align with existing literature on the challenges of sustaining effective malaria control in resource-constrained settings where health infrastructure is limited and prevention tools are inconsistently distributed [10,11]. In addition, CHWs identified a growing burden of non-communicable diseases such as hypertension, diabetes, and cervical cancer. Yet, access to screening and early diagnosis remains minimal, and public awareness is limited. These trends echo broader shifts in disease burden observed across sub-Saharan Africa, where fragile health systems are now grappling with a dual burden of infectious and chronic diseases [12].

The study underscores the profound impact of poverty, distance, and inadequate infrastructure on health-seeking behaviors. CHWs reported that many community members delay care due to long travel distances, lack of transportation, and the unaffordability of private services. Even when patients reach health centers, they often encounter stockouts, overburdened staff, and fragmented service delivery. These barriers are well-documented in rural health systems globally and reflect longstanding health equity challenges that disproportionately affect underserved communities [5,13,14]. Additionally, the mismatch between community knowledge and actual health practices, particularly in nutrition, preventive care, and water sanitation, highlights the limitations of information-based interventions in the absence of structural change. CHWs noted that economic constraints, seasonal food shortages, and inadequate service coverage prevent families from acting on health knowledge, reinforcing the importance of addressing upstream social determinants.

A central theme emerging from this study is the dual burden carried by CHWs: they are deeply embedded in their communities and highly trusted, yet they operate with limited tools, support, or recognition. CHWs are often the first point of contact for health concerns, particularly in remote areas, but they are not equipped with the diagnostic tools, medicines, or transportation needed to respond effectively. The erosion of community trust due to these limitations was noted repeatedly. Nonetheless, CHWs demonstrated resilience, adaptability, and a strong commitment to serving their communities. They expressed a desire for greater training, meaningful supervision, and inclusion in decision-making, factors known to contribute to CHW motivation, retention, and program sustainability [15,16]. Their recommendations reflect both practical resource needs and a call for systemic inclusion, suggesting that effective CHW programming must go beyond task allocation to include workforce empowerment, partnership, and recognition.

The findings of this study have important implications for community health program design. First, health system strengthening efforts should prioritize reliable supply chains for diagnostics and essential medicines, particularly in high-burden settings like Pader District. Second, investments in transportation, such as bicycles or travel stipends, can significantly enhance CHW reach and effectiveness in geographically dispersed communities. Third, digital tools offer an opportunity to improve CHW data management and referral systems, provided they are accompanied by training and support. Equally important are non-material interventions: regular supportive supervision, recognition of CHW contributions, and meaningful inclusion in planning and feedback mechanisms. These strategies have been shown to enhance CHW performance and foster a sense of ownership and accountability [17]. Finally, the findings point to the need for integrated programming that addresses both infectious and non-communicable diseases, bridges health literacy with structural support, and builds trust between communities and health systems.

This study offers several important strengths. First, it captures the voices and lived experiences of CHWs across a range of settings in Pader District, providing a nuanced understanding of health challenges and systemic barriers from the perspective of frontline providers. The use of focus group discussions enabled rich dialogue, allowing participants to build on one another’s insights and highlight shared concerns and priorities. Second, the study was designed and implemented in partnership with local collaborators, enhancing cultural relevance, contextual sensitivity, and ethical rigor. Finally, the integration of community perspectives into the analysis and interpretation of findings supports the development of grounded, actionable recommendations to inform program improvement and policy at the district level and beyond.

In addition to its strengths, this study has several limitations. Findings are specific to Pader District and may not be generalizable to all regions of Uganda. Focus group dynamics may have influenced participant responses, with potential for social desirability bias. Additionally, while CHWs are deeply knowledgeable about community health, their perspectives may not capture the full range of community experiences, particularly among groups less engaged with the CHW system.

## Conclusion

Community health workers in Pader District play a vital role in addressing persistent health challenges in rural Northern Uganda. Their experiences reveal a high burden of both infectious and non-communicable diseases, alongside structural barriers such as limited access to care, medicine stockouts, and inadequate infrastructure. Despite these obstacles, CHWs remain trusted figures in their communities and serve as essential connectors between households and the formal health system. Strengthening CHW programs requires not only the provision of basic tools, transportation, and regular training, but also supportive supervision, consistent stipends, and symbolic recognition. Equally important is the inclusion of CHWs in program planning and decision-making, ensuring that interventions are grounded in local realities. Future directions should focus on implementing and evaluating scalable interventions that respond directly to CHW-identified needs like mobile reporting tools, improved supply chains, and peer mentorship networks. Further research should examine the impact of these changes on CHW motivation, service quality, and health outcomes. Policymakers and implementing partners must prioritize CHW voices and leadership to build resilient, community-centered health systems in Uganda and similar settings.

## Data Availability

Data and manuscript draft are available through open science framework. Ebbs, D. (2025, September 6). Exploring Local Health Knowledge and Access: Focus Group Findings from Community Health Workers in Pader District, Uganda. Retrieved from osf.io/d2jgr

## Acknowledgements

I would like to sincerely thank Drs Ebbs and Bongomin, who graciously invited me to join their research team and connected me to incredible collaborators in Uganda, for their invaluable guidance and feedback throughout this project. I would also like to extend much gratitude to DHO Dr. Oyoo Benson, Bosco, David, Jacob, as well as the rest of the NUMEM team and my friends in Pader, none of this work would have been possible without their important input and hospitality every day.

